# External Validation of Predictive Models for Diagnosis, Management and Severity of Pediatric Appendicitis

**DOI:** 10.1101/2024.10.28.24316300

**Authors:** Ričards Marcinkevičs, Kacper Sokol, Akhil Paulraj, Melinda A. Hilbert, Vivien Rimili, Sven Wellmann, Christian Knorr, Bertram Reingruber, Julia E. Vogt, Patricia Reis Wolfertstetter

## Abstract

**Background:** Appendicitis is a common condition among children and adolescents. Machine learning models can offer much-needed tools for improved diagnosis, severity assessment and management guidance for pediatric appendicitis. However, to be adopted in practice, such systems must be reliable, safe and robust across various medical contexts, e.g., hospitals with distinct clinical practices and patient populations.

**Methods:** We performed external validation of models predicting the diagnosis, management and severity of pediatric appendicitis. Trained on a cohort of 430 patients admitted to the Children’s Hospital St. Hedwig (Regensburg, Germany), the models were validated on an independent cohort of 301 patients from the Florence-Nightingale-Hospital (Düsseldorf, Germany). The data included demographic, clinical, scoring, laboratory and ultrasound parameters. In addition, we explored the benefits of model retraining and inspected variable importance.

**Results:** The distributions of most parameters differed between the datasets. Consequently, we saw a decrease in predictive performance for diagnosis, management and severity across most metrics. After retraining with a portion of external data, we observed gains in performance, which, nonetheless, remained lower than in the original study. Notably, the most important variables were consistent across the datasets.

**Conclusions:** While the performance of transferred models was satisfactory, it remained lower than on the original data. This study demonstrates challenges in transferring models between hospitals, especially when clinical practice and demographics differ or in the presence of externalities such as pandemics. We also highlight the limitations of retraining as a potential remedy since it could not restore predictive performance to the initial level.

## 1 Introduction

Acute appendicitis is a common condition among children and adolescents treated in pediatric surgery departments due to abdominal pain [8, 29]. Diagnosis relies on clinical signs and symptoms (in particular, their dynamics and progression under close observation), laboratory tests and imaging, whereas postoperative classification is based on intraoperative findings and histology [13]. Scoring systems, such as Alvarado Score (AS) and Pediatric Appendicitis Score (PAS), can facilitate clinical assessment [14, 15]. The classical treatment of pediatric appendicitis is surgical, although conservative treatment with antibiotics can be an option in certain cases [11, 34, 35]. Additionally, spontaneous resolution of uncomplicated appendicitis has been observed and reported, which supports antibiotic-free management based on supportive care for qualifying cases [23, 24, 27].

Despite new developments and technologies, early and accurate detection, preoperative classification, and treatment strategy selection are still challenging, especially in young children [5, 8, 9]. Widely used clinical and laboratory parameters alone are mostly non-specific at identifying appendicitis [1, 39]. Imaging modalities are important tools to guide management and avoid negative appendectomies, but they have limitations, such as operator (investigator) dependency on ultrasonography, radiation exposure for computed tomography, and availability and feasibility of magnetic resonance imaging, not to mention the costs [13, 19].

Recent years have marked impressive progress in Machine Learning (ML) research and the increasing proliferation of tools built upon this technology in medicine. ML algorithms promise to aid in the detection, management and treatment of various diseases, thus improving the overall quality and effectiveness of healthcare. In relation to pediatric appendicitis, ML has been used to diagnose and manage patients suspected of developing this condition [2–4, 12, 18, 20, 21, 26, 28, 33, 38]; specifically, such tools were developed to predict diagnosis, management and severity of pediatric appendicitis. These models either rely exclusively on standard clinical and laboratory data [2, 4, 12, 18, 38], or additionally utilize imaging modalities (obtained through various methods, e.g., computed tomography or ultrasonography) either directly in their raw format or by extracting hand-crafted annotations [3, 20, 21, 26, 28, 33].

Although promising and practical, ML-based tools for pediatric appendicitis are rarely deployed in practice due to the translational barrier inherent to medical machine learning research [32]. To overcome this challenge, predictive models need to be validated on external datasets and later go through rigorous clinical trials (which tend to be complex, time-consuming and costly) [37]. In this study we make a step in this direction and follow up on our previous work where we developed ML models [21] for predicting diagnosis (*appendicitis* vs. *no appendicitis*), treatment assignment (*surgical* vs. *conservative*) and complications (*complicated appendicitis* vs. *uncomplicated* or *no appendicitis*) of pediatric appendicitis. Specifically, we conduct a principled external validation of the aforementioned ML tools on tabular electronic health records collected in a different hospital, exploring potential challenges associated with the transfer of our predictive models.

The original models (logistic regression, random forest and gradient boosting, all achieving strong performance) were developed on a dataset of 430 patients aged 0 to 18 years admitted with abdominal pain and suspected appendicitis to the Department of Pediatric Surgery at the tertiary Children’s Hospital St. Hedwig in Regensburg, Germany, over the period of 2016–2018 [3, 21]. The original dataset consists of demographic, clinical, scoring, laboratory and ultrasound (US) predictor variables (see Table 1 for their list).^1^ The external validation dataset was acquired at the Department of Pediatric Surgery and Pediatric Traumatology, Florence-Nightingale-Hospital, Düsseldorf, Germany. This cohort consists of 301 pediatric patients hospitalized between 2015 and 2022, and the dataset format and predictor variables adhere to the format of the Regensburg dataset. The study design is summarized schematically in Figure 1.

**Table 1.** Description of the Regensburg and Düsseldorf datasets containing summary statistics for each variable. For numerical variables, we report medians alongside interquartile ranges; categorical variables are binarized and summarized as frequencies. Additionally, we report adjusted *p*-values from the unpaired two-sample *t*-test and chi-squared test for proportions. For significant differences, *p*-values are given in **bold**.

| Feature | | Regensburg<br>$n = 430$ | Düsseldorf<br>$n = 301$ | $p$-value |
| --- | --- | --- | --- | --- |
| **Demographic** | Age [years] | 11.5 [9.3, 13.9] | 10.1 [7.7, 11.7] | $\leq \mathbf{0.001}$ |
|  | Male sex [%] | 53.7 | 58.1 | 0.260 |
| | Height [cm] | 150.5 [138.0, 162.9] | 140.0 [128.3, 150.0] | $\leq \mathbf{0.001}$ |
| | Weight [kg] | 42.0 [31.1, 55.0] | 35.0 [26.0, 43.0] | $\leq \mathbf{0.001}$ |
| | Body mass index [kg/m<sup>2</sup>] | 18.1 [15.85, 21.2] | 17.9 [15.8, 20.1] | $\leq \mathbf{0.050}$ |
| **Scoring** | Alvarado score [points] | 6.0 [4.0, 7.0] | 6.0 [5.0, 7.0] | 0.054 |
| | Pediatric appendicitis score [points] | 5.0 [4.0, 6.0] | 6.0 [5.0, 7.0] | $\leq \mathbf{0.001}$ |
| **Clinical** | Peritonitis [%] | 38.4 | 64.1 | $\leq \mathbf{0.001}$ |
| | Migration of pain [%] | 25.6 | 46.6 | $\leq \mathbf{0.001}$ |
|  | Tenderness in right lower quadrant [%] | 97.0 | 94.6 | 0.129 |
| | Rebound tenderness [%] | 34.4 | 43.0 | $\leq \mathbf{0.050}$ |
| | Cough tenderness [%] | 27.0 | 48.3 | $\leq \mathbf{0.001}$ |
| | Psoas sign [%] | 30.5 | 41.8 | $\leq \mathbf{0.010}$ |
| | Nauseous/vomitting [%] | 56.3 | 68.1 | $\leq \mathbf{0.010}$ |
| | Anorexia [%] | 29.1 | 68.1 | $\leq \mathbf{0.001}$ |
| | Body temperature [°C] | 37.4 [37.0, 38.2] | 37.0 [36.5, 37.7] | $\leq \mathbf{0.001}$ |
|  | Dysuria [%] | 5.4 | 4.0 | 0.415 |
| | Abnormal stool [%] | 27.8 | 19.5 | $\leq \mathbf{0.050}$ |
| **Laboratory** | White blood cell count [$10^3/\mu\text{l}$] | 11.9 [8.4, 15.8] | 14.9 [10.3, 19.3] | $\leq \mathbf{0.001}$ |
|  | Neutrophils [%] | 74.9 [59.1, 82.9] | 72.7 [59.8, 82.1] | 0.293 |
| | C-reactive protein [mg/l] | 7.0 [1.0, 31.3] | 19.0 [5.0, 58.0] | $\leq \mathbf{0.010}$ |
| | Ketones in urine [%] | 38.4 | 53.7 | $\leq \mathbf{0.001}$ |
| | Erythrocytes in urine [%] | 22.1 | 33.9 | $\leq \mathbf{0.010}$ |
|  | White blood cells in urine [%] | 12.4 | 17.0 | 0.153 |
| **Ultrasound** | Visibility of appendix [%] | 64.5 | 24.3 | $\leq \mathbf{0.001}$ |
| | Appendix diameter [mm] | 7.3 [6.0, 9.1] | 9.0 [7.0, 12.0] | $\leq \mathbf{0.001}$ |
| | Free intraperitoneal fluid [%] | 43.6 | 25.2 | $\leq \mathbf{0.001}$ |
| | Irregular appendix layers [%] | 35.9 | 7.2 | $\leq \mathbf{0.001}$ |
| | Target sign [%] | 46.0 | 30.8 | $\leq \mathbf{0.010}$ |
|  | Appendix perfusion [%] | 65.5 | — | — |
| | Surrounding tissue reaction [%] | 71.7 | 16.4 | $\leq \mathbf{0.001}$ |
| | Pathological lymph nodes [%] | 68.5 | 2.7 | $\leq \mathbf{0.001}$ |
| | Mesenteric lymphadenitis [%] | 80.4 | 6.6 | $\leq \mathbf{0.001}$ |
| | Thickening of the bowel wall [%] | 40.9 | 11.9 | $\leq \mathbf{0.001}$ |
| | Ileus [%] | 14.5 | 0.0 | $\leq \mathbf{0.001}$ |
| | Coprostasis [%] | 37.8 | 4.8 | $\leq \mathbf{0.001}$ |
| | Meteorism [%] | 72.9 | 26.4 | $\leq \mathbf{0.001}$ |
| | Enteritis [%] | 46.3 | 0.0 | $\leq \mathbf{0.001}$ |
| **Response** | Appendicitis [%] | 57.2 | 76.2 | $\leq \mathbf{0.001}$ |
| | Surgical management [%] | 38.4 | 80.5 | $\leq \mathbf{0.001}$ |
|  | Complicated appendicitis [%] | 11.9 | 16.4 | 0.091 |

**Fig. 1.**
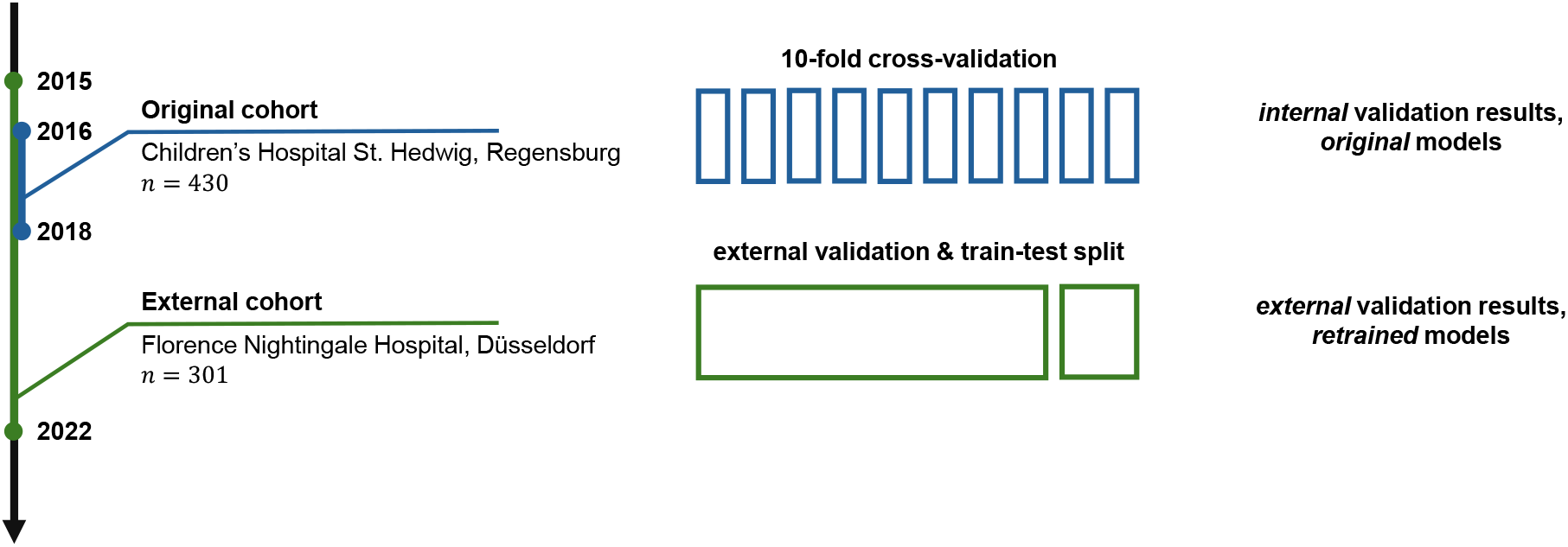
An overview of the study design. The original predictive models were trained and validated on the cohort of patients (*n* = 430) from Regensburg, Germany [21]. This article presents the results of the external validation on another cohort (*n* = 301) from Düsseldorf, Germany. In particular, in this study, we validate the original models on the external data and retrain them to assess the potential for improvement.

In this retrospective study, we present an external validation of the aforementioned models on a new and independent cohort of patients. To this end, we:

1. compare the datasets to better understand their differences (Section 3.1);
2. evaluate the models without any adaptation to test their *external validity* under real-world distribution shift (Section 3.2);
3. retrain the models, and then evaluate and compare them again to explore possible gains in performance (Section 3.2); and
4. study feature importance across the models to elucidate their functioning (Section 3.3).

Our study demonstrates the transferability of the models across hospitals and outlines the steps necessary to facilitate such a safe adaptation.

## 2 Material and Methods

### 2.1 External Data Acquisition and Description

To facilitate external validation, we collected and reviewed retrospective data from children and adolescents aged 0–17 years who were admitted to the Department of Pediatric Surgery and Pediatric Traumatology at Florence-Nightingale-Hospital in Düsseldorf with abdominal pain and suspected appendicitis from January 1st, 2015 to February 1st, 2022. Patients who had undergone an appendectomy before their admission were excluded. Similarly, we did not include subjects with chronic intestinal diseases or current antibiotic treatment if therapy was conservative. In total, 301 patients met the inclusion criteria. The study, including data acquisition and transfer, was approved by the Ethics Committee of the University of Regensburg (18-1063-101, 18-1063-3-101) and was performed in accordance with the relevant guidelines and regulations.

In terms of management, the cohort was divided into conservative and operative groups. Patients admitted and receiving supporting therapy, e.g., intravenous fluids, enemas and analgesics, with clinical improvement without surgery were classified as conservative. Otherwise, having undergone an appendectomy, subjects were labeled as operative. For the surgical group, histological findings were recorded. In the case of negative appendectomy (i.e., histology presenting a normal appendix without inflammation), the corresponding data were used to predict the diagnosis and severity but not the management.

As in the prior study [21], diagnosis (*appendicitis* vs. *no appendicitis*) was assessed for all included patients. For patients treated surgically, appendicitis diagnosis was based on histology. In the nonoperative group, patients were classified retrospectively as having appendicitis if their AS or PAS were at least 4, combined with an appendix diameter of ≥ 6 mm. Conservatively treated patients classified as having appendicitis were followed up after discharge for recurrences. Patients who had a recurrence and underwent secondary operation were relabeled as surgical in the analysis. The follow-up telephone interview was performed at least one year after discharge, between January 2023 and February 2024. Informed consent was obtained from the parents or legal representatives of the patients who underwent the follow-up.

Furthermore, appendicitis severity was assessed. Patients treated non-operatively, both with and without appendicitis, with no recurrence during the follow-up period were classified as *uncomplicated*. For patients treated operatively, classification was based on the histology: *simple/uncomplicated* (subacute/catharral/chronic, phlegmonous) or *complicated* (abscess, perforation).

During the exploratory analysis presented below, we compute summary statistics across both datasets – the original from Regensburg and the external from Düsseldorf – and perform hypothesis tests for the differences between internal and external data. Specifically, we report medians and interquartile ranges (IQR) for numerical attributes and frequencies for categorical features. For statistical analysis, we utilize the unpaired two-sample *t*-test and Pearson’s chi-squared test for the equality of proportions. We adjust the resulting *p*-values for multiple comparisons to control the false discovery rate using the Benjamini-Hochberg procedure [6] at the level *q* = 0.05.

### 2.2 Original Predictive Models

We leverage the dataset from the Florence-Nightingale-Hospital, Düsseldorf, for the external validation of the predictive models developed on the Regensburg cohort [21]. The original analysis [21] was concerned with predicting three response variables: (i) diagnosis (*appendicitis* vs. *no appendicitis*), (ii) management (*surgical* vs. *conservative*), and (iii) severity (*complicated appendicitis* vs. *uncomplicated* or *no appendicitis*). In particular, logistic regression (LR), random forest (RF) [10] and gradient boosting (GB) [16] models were trained on the dataset of 430 patients with 38 predictor variables.

In the current study, we train these models on the *full* Regensburg cohort, replicating the original R programming language code [21, 25] in the Python programming language (v3.11.9) using the scikit-learn library (v1.4.2). We use hyperparameter configurations and perform preprocessing steps similar to those described in the original study [21], imputing missing values with the *k*-nearest neighbors algorithm (with *k* = 5). Note that we limit our analysis to models trained on the full set of features and we do not consider ablations with feature selection or without the US-related variables.

### 2.3 Model Retraining

In addition to the purely external validation, we retrain the predictive models on a combination of the Regensburg and Düsseldorf cohorts, building the models on the 100% of the Regensburg and 80% of the Düsseldorf data. In this setting, we test the models on the remaining, withheld 20% of the external dataset (the data were split at random). We conduct this experiment to gauge the possible benefits of a multicenter cohort approach and to better understand if the predictive performance improves with the inclusion of external data points in the training set.

### 2.4 Evaluation

For both original and retrained model evaluation, we report the area under the receiver operating characteristic (AUROC) and precision-recall (AUPR) curves. Additionally, we investigate the tradeoffs among sensitivity, specificity as well as positive (PPV) and negative (NPV) predictive values by varying the threshold applied to the classifiers’ output. Lastly, to better understand the models’ predictions, we compute the permutation feature importance [10] of predictor variables using the test set.

## 3 Results

### 3.1 External Dataset

Both of the datasets investigated in this study are overviewed in Table 1. Therein we report summary statistics for all the variables observed across the Regensburg (*n* = 430) and Düsseldorf (*n* = 301) cohorts. Additionally, we provide the adjusted results of statistical hypothesis tests for the differences in means and proportions of values respectively for numerical and categorical variables.

We observe significant differences across the distribution of most variables. Generally, subjects from the Düsseldorf cohort are younger and exhibit a higher frequency of clinical examination findings. Similarly, the external data exhibit overall higher laboratory parameter values for the variables correlated with appendicitis. Despite this, we observe no statistically significant difference in the neutrophil percentage, likely due to the high rate of missing values for this predictor in the external dataset (see Figure 2).

**Fig. 2.**
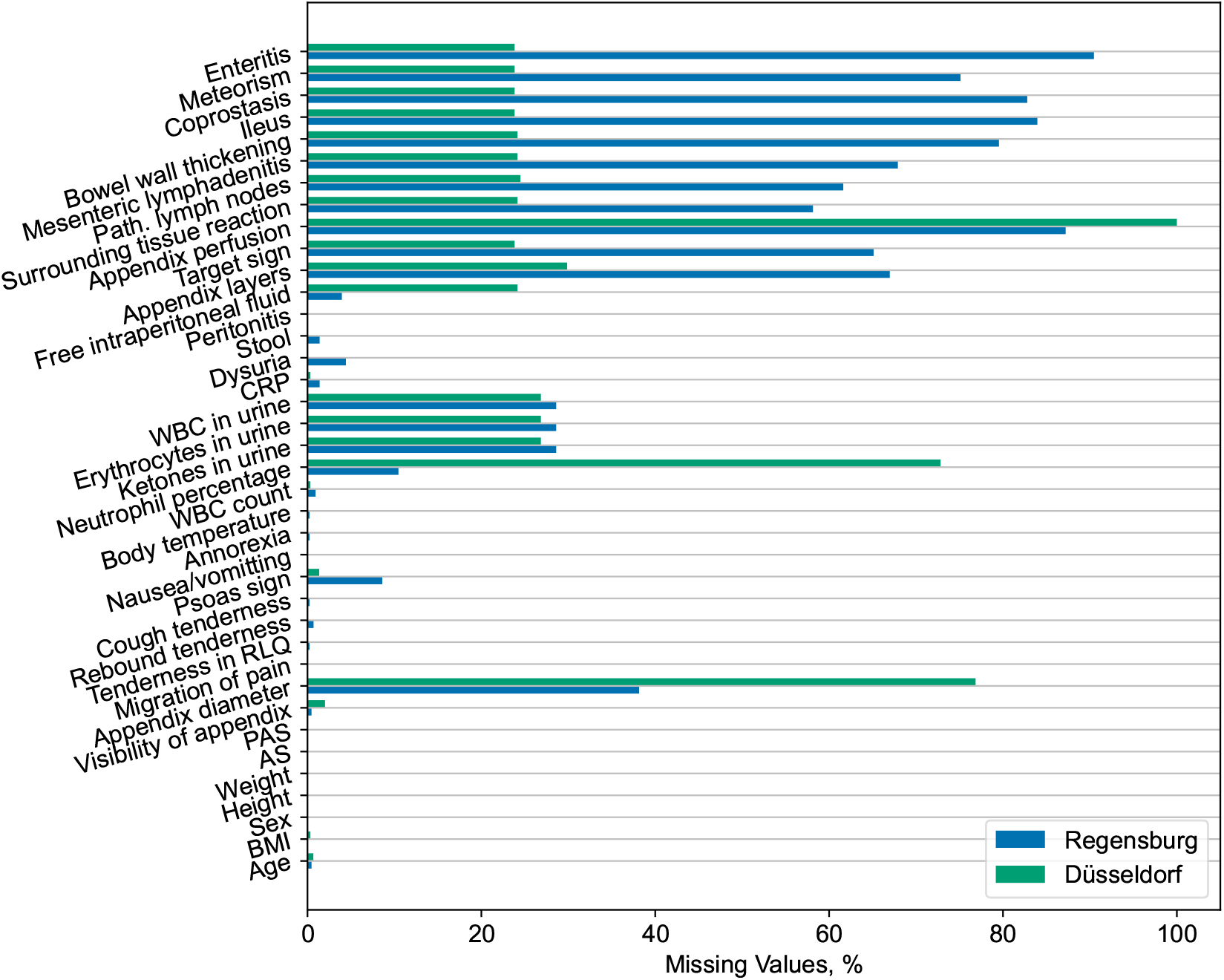
Percentages of missing values across all features for the original Regensburg and external Düsseldorf data. We observe considerable differences in the rates of missing values, especially for the ultrasonographic findings and neutrophil percentage.

Furthermore, the Düsseldorf cohort has a lower frequency of positive US findings. We attribute this trend to the higher rate of missing values for relevant variables in the Regensburg dataset (refer to Figure 2) and the fact that the summary statistics shown in Table 1 have been calculated only across the non-missing entries without imputation. By contrast, the reported appendix diameter is significantly larger for the external dataset subjects. Lastly, it is worth noting that the information about the appendix perfusion is entirely missing in the Düsseldorf dataset.

The datasets also differ considerably in two of the response variables: diagnosis and management. The Düsseldorf cohort has a significantly higher prevalence of appendicitis cases (76.2% vs. 57.2%) with, consequently, more patients managed surgically (80.5% vs. 38.4%). While the external dataset has a higher prevalence of complicated appendicitis cases (16.4% vs. 11.9%), this difference is not statistically significant.

In summary, the external dataset from Düsseldorf and the original dataset from Regensburg exhibit statistically significant differences with regard to the distribution of the majority of the observed variables (consult Table 1), including the response variables. Moreover, the frequency of missing values also varies across the cohorts (refer to Figure 2). These dissimilarities potentially pose challenges for the generalization of predictive models across institutions.

### 3.2 Predictive Performance

We now turn to the external validation of the ML models. Table 2 contains AUROC and AUPR measurements for predicting the diagnosis, management and severity of appendicitis on the Regensburg and Düsseldorf datasets. The results for the Regensburg cohort are taken from the original work [21] and were obtained by 10-fold cross-validation. When validating on the Düsseldorf data, we assess the variability in performance using bootstrapping. For reference, we additionally include the expected metric values for a fair coin flip (*random*), which serve as our baselines.

**Table 2.** Validation results for the logistic regression (LR), random forest (RF) and gradient boosting (GB) models predicting the diagnosis, management and severity of appendicitis. The results on the Regensburg dataset are copied from the original study [21], which conducted 10-fold cross-validation. For the Düsseldorf data, we report averages and standard deviations obtained by bootstrapping for the models trained exclusively on the Regensburg cohort (*original*) and retrained on both cohorts (*retrained*). The predictive performance is assessed with the areas under the receiver operating characteristic (AUROC) and precision-recall (AUPR) curves.

| Dataset | Model | Diagnosis |  | Management |  | Severity |  |
| --- | --- | --- | --- | --- | --- | --- | --- |
|  |  | AUROC | AUPR | AUROC | AUPR | AUROC | AUPR |
| Regensburg | Random [21] | 0.50 | 0.43 | 0.50 | 0.38 | 0.50 | 0.12 |
|  | Original LR [21] | 0.91±0.04 | 0.88±0.07 | 0.90±0.04 | 0.88±0.06 | 0.82±0.13 | 0.53±0.26 |
|  | Original RF [21] | 0.96±0.01 | 0.94±0.03 | 0.94±0.02 | 0.92±0.05 | 0.90±0.08 | 0.70±0.17 |
|  | Original GBM [21] | 0.96±0.01 | 0.94±0.03 | 0.94±0.02 | 0.93±0.04 | 0.90±0.07 | 0.64±0.21 |
| Düsseldorf | Random | 0.50 | 0.76 | 0.50 | 0.81 | 0.50 | 0.16 |
|  | Original LR | 0.80±0.04 | 0.92±0.02 | 0.80±0.04 | 0.94±0.02 | 0.70±0.06 | 0.34±0.08 |
|  | Original RF | 0.85±0.03 | 0.95±0.01 | 0.85±0.03 | 0.96±0.01 | 0.75±0.04 | 0.45±0.07 |
|  | Original GBM | 0.83±0.03 | 0.94±0.02 | 0.82±0.03 | 0.95±0.01 | 0.72±0.04 | 0.40±0.07 |
|  | Retrained LR | 0.84±0.08 | 0.94±0.04 | 0.83±0.08 | 0.95±0.03 | 0.74±0.12 | 0.45±0.17 |
|  | Retrained RF | 0.87±0.06 | 0.96±0.02 | 0.83±0.08 | 0.95±0.03 | 0.75±0.11 | 0.49±0.17 |
|  | Retrained GBM | 0.86±0.07 | 0.95±0.03 | 0.82±0.09 | 0.95±0.03 | 0.75±0.11 | 0.47±0.18 |

For the models trained exclusively on the Regensburg data (*original*), we observe a sizable decrease in the average AUROC for the diagnosis and management when evaluating on the external dataset. For example, the AUROC of the random forest model decreases from 96% to 85% for the diagnosis and from 94% to 85% for the management. In contrast the external AUPR is comparable to the one from the internal validation for these response variables. For the severity, we observe a larger overall decrease in both metrics. For instance, for the random forest, the AUROC decreases from 90% to 75%, and the AUPR drops from 70% to 45%.

Additionally, we explore the tradeoff between the sensitivity, specificity, PPV and NPV while varying the value of the threshold applied to the classifiers’ output. We focus our analysis exclusively on the random forest model as it exhibits the most balanced performance across all the response variables for both datasets. These findings are summarized in Figure 3. For the diagnosis and management targets, using the threshold value of 0.50 explored in the original analysis [21], we observe a deterioration in the classifiers’ sensitivity, specificity and NPV. For the severity target, by contrast, there is a decline in sensitivity and PPV. Arguably, these changes may be related to the prevalence shift [17] described in Section 3.1 and suggest the necessity for the threshold and model recalibration.

**Fig. 3.**
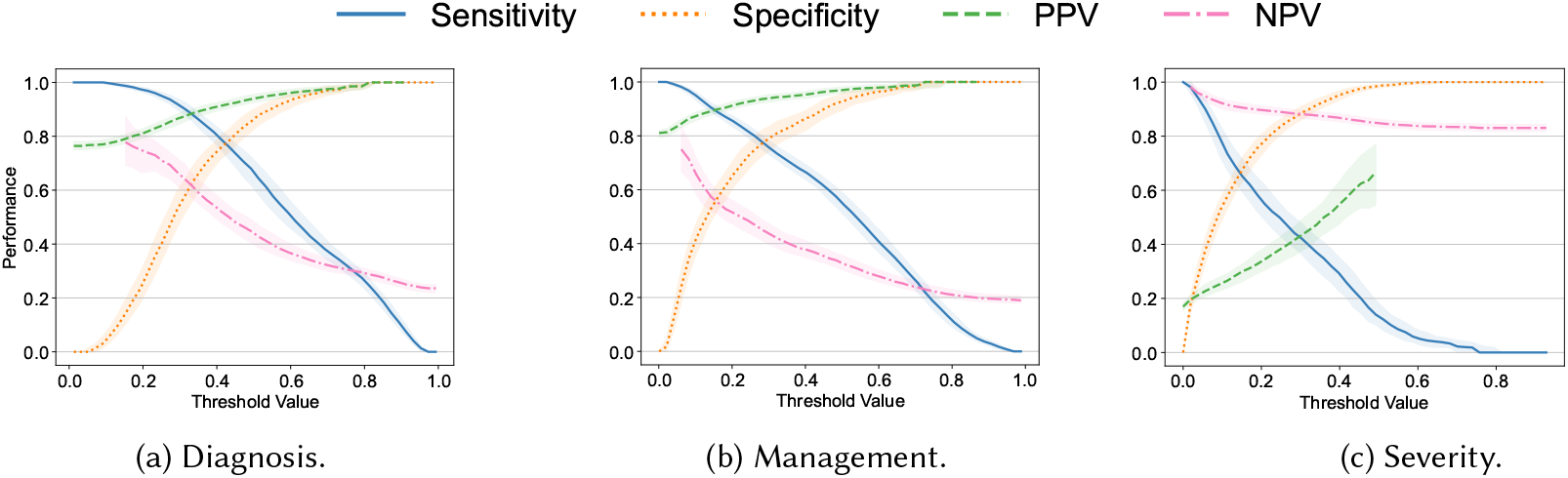
Sensitivity, specificity as well as positive (PPV) and negative (NPV) predictive values plotted against the value of the threshold applied to the output of the random forest model trained exclusively on the original Regensburg dataset for the (a) diagnosis, (b) management and (c) severity of appendicitis. All the metrics were assessed on the external (Düsseldorf) dataset. Bold lines correspond to the medians with the confidence bounds given by the interquartile ranges.

To verify if the models’ performance improves after including a portion of the Düsseldorf data in the training set, we retrain all the models on the aforementioned mixture of the Regensburg and Düsseldorf subjects (see the *retrained* models in Table 2), assessing them on the withheld portion of the external dataset. For all three classifiers, the average AUROC and AUPR metric values attained on the Düsseldorf data increase moderately after retraining. However, the resulting level of performance is still substantially lower than that of the original models on the Regensburg dataset. The lack of bigger improvement in predictive performance may be due to distribution shift, in particular the discrepancies in the missingness patterns and reporting across the two datasets (see Figure 2).

### 3.3 Feature Importance

To elucidate the predictions made by our models on the external dataset, we calculate *permutation feature importance* on the test set. Specifically, we assess the importance of individual predictors by permuting (i.e., shuffling) their values and then quantifying the resulting change in the AUROC metric. The outcomes of this analysis are summarized in Figure 4. We limit our investigation to the diagnosis response variable and the random forest model given that it attained the best well-balanced performance across all the settings (refer back to Table 2).

**Fig. 4.**
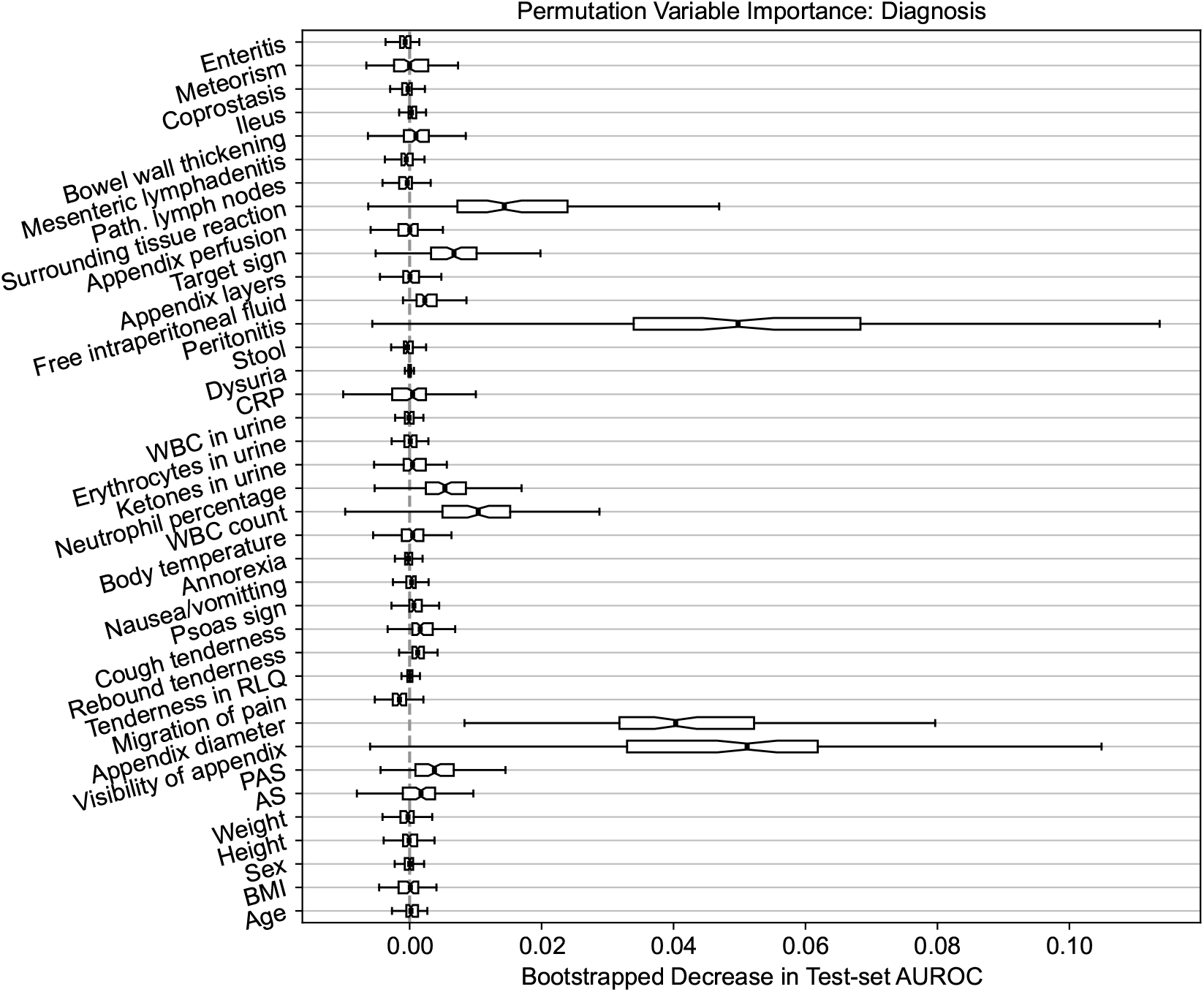
Permutation feature importance for the random forest model predicting the diagnosis of appendicitis. The importance is quantified by the decrease in the AUROC predictive performance metric after permuting the values of the predictor variable of interest. The variability in importance is assessed using bootstrapping and it is visualized using box plots.

Similar to the original findings on the Regensburg data [21], the three most important features are the diameter and visibility of the appendix as well as peritonitis. Likewise, the surrounding tissue reaction, target sign, WBC count and neutrophil percentage have an importance score, on average, above 0. Generally, the variable importance on the Düsseldorf data follows a pattern comparable to the results obtained previously on the Regensburg cohort. However, the variability across bootstrap resamples is considerably higher. Nonetheless, these results are not indicative of any concerning trends or spurious associations and fall well within our expectations. Notably, these observations hold for the other two response variables; for treatment the three most important features are peritonitis, appendix diameter and WBC count, and for complications these are CRP, peritonitis and appendix diameter, which is consistent with the results reported in the Regensburg study [21]. In the original analysis, the most important predictors were appendix diameter, peritonitis and CRP respectively for diagnosis, treatment and complications.

## 4 Discussion

In this article, we performed a comprehensive external validation of ML models for predicting the diagnosis, management and severity in pediatric patients with suspected appendicitis (see Figure 1). Specifically, we have focused on the models initially trained on the dataset from the tertiary care hospital in Regensburg, Germany [21]. To conduct the analysis, we have acquired an external dataset at the Florence-Nightingale-Hospital in Düsseldorf, Germany.

We observed that the external Düsseldorf dataset presents a statistically significant shift in the distribution of the covariates (captured in Table 1), including the response variables. Furthermore, the rates of missing values differ considerably across the two hospitals (as shown in Figure 2), especially for US-related variables and the percentage of neutrophils. Such discrepancies pose substantial challenges to the transferability of ML models to settings different from those considered at the training time [36].

In assessing the models’ predictive performance (reported in Table 2), we observed the patients’ diagnoses and treatment assignments could be predicted on the external Düsseldorf data by the models trained solely on the Regensburg cohort. In particular, compared to the original analysis [21], there was no decrease in AUPR and a moderate 10 percentage point decrease in AUROC. These performance levels are close to the AUROC of 90% reported as the baseline in a recent systematic review assessing the accuracy of artificial intelligence-based tools used in pediatric appendicitis diagnosis [30]. In contrast, the predictive performance for the severity decreased more substantially. In addition to AUROC and AUPR measurements, we examined the tradeoff among the sensitivity, specificity, PPV and NPV (visualized in Figure 3). Furthermore, the feature importance analysis on the external dataset (shown in Figure 4) exhibited no concerning patterns.

Several risk prediction models for appendicitis have been built, evaluated and validated, including for varied patient cohorts (adults vs. children), in- and out-patient treatments, and outcomes. A recent prospective multicenter study also evaluated the performance of risk scores to identify appendicitis among children brought to the hospital emergency department [22]. It identified that low appendicitis scores (*≤* 2 for Alvarado or PAS, or *≤* 3 for Shera-Score) can be used to preselect children who can be discharged without further evaluation, but was unable to offer guidelines to select children who should proceed directly to surgery, indicating that patients with medium and high risk scores should undergo routine imaging examination [22]. Our original and current study relied on a wide selection of variables, including the aforementioned risk scores (Alvarado score and PAS) and imaging parameters (ultrasound) from pediatric patients who were suspected of appendicitis, not only to exclude children with a low probability of appendicitis, but also to predict the diagnosis, management and severity of appendicitis [27].

To explore the potential of model updating [36], we retrained the classifiers on a mixture of the two datasets. This led to a moderate increase in AUROC and AUPR across all the target variables (refer to Table 2), suggesting that model updating, indeed, helps to tackle cross-hospital distribution shifts. More generally, our empirical findings indicate some degree of transferability of the considered predictive models across the two hospitals. Nonetheless, the decrease in predictive performance across several evaluation metrics is noticeable and could not be fully mitigated by retraining alone (as demonstrated by Table 2 and Figure 3). We hypothesize that this decrease in performance may be attributed to the shift in the prevalence of appendicitis cases, different missing value and data recording patterns, and variability in patient management routines. Below, we discuss these challenges in more detail.

As stated in Section 3.1, the distribution of most parameters differed across the two datasets. Unique regional and internal hospital practices can, at least partially, explain the observed differences. Notably, the dataset from Regensburg was acquired from in-hospital patients admitted to a pediatric surgery department of a *specialized* pediatric hospital. The Düsseldorf dataset, on the other hand, was acquired from a pediatric surgery department of a *general* hospital with other surgical disciplines, such as general and orthopedic surgery. As a consequence, children aged 14 years or older were treated by general surgeons in Düsseldorf, and only those younger than 14 were seen and managed by pediatric surgeons. Consequently, only the latter group of patients was included in the study, which explains why the children from the Florence-Nightingale-Hospital were younger (median and IQR in years: 10.1 [7.7, 11.7]) than in Regensburg (11.5 [9.3, 13.9]).

The differences in the frequency of variable documentation reflect the internal organizational habits of the hospitals and departments, including variations in standardized admission reports and internal emergency department standards. Additionally, in Regensburg, children and adolescents were admitted by pediatric surgeons or residents in pediatric surgery or pediatrics, whereas in Düsseldorf, the admission was performed by both pediatric surgeons or residents and residents in general or orthopedic surgery working at the emergency department. Consequently, the ultrasound performance and report documentation differs across the two datasets.

Additionally, the clinical pathways and referral practices differed between the two institutions. In Düsseldorf, pediatricians and general practitioners were more likely to refer children to pediatric surgery when the diagnosis of appendicitis was already relatively clear. This preselection process likely contributed to the higher prevalence of confirmed appendicitis and surgical treatment cases in the Düsseldorf cohort. In contrast, the specialized pediatric setting in Regensburg enabled children with less specific abdominal symptoms to be evaluated by multiple specialties, such as pediatric gastroenterology, leading to a broader spectrum of differential diagnoses and, in some cases, more conservative management. Furthermore, the clinical pathway in Regensburg often involved admitting children with tenderness in the right iliac fossa to pediatric surgery for further observation, even in cases with unclear diagnosis. These patients were included in the cohort and may partly explain the higher rate of non-appendicitis cases, lower CRP values and lower frequency of surgical intervention.

Another noteworthy aspect is the time period of data acquisition. While the cohort from Regensburg included patients from January 2016 to December 2018, the Düsseldorf data were acquired from January 2015 to February 2022. Therefore, the latter cohort also included patients admitted during the COVID-19 pandemic and post-pandemic individuals, and negative appendectomy rates were lower during the pandemic [7] as patients might have sought medical care or have been referred to the hospital only if the positive diagnosis had been deemed more probable. This factor, alongside the higher frequency of delayed hospital presentations, might have contributed to the higher appendicitis prevalence and the higher rate of complicated cases observed in Düsseldorf [7, 31].

From the medical perspective, the limitations of the current study are similar to those reported in the original work that developed ML models on the Regensburg cohort [21]. These include absence of confirmed appendicitis diagnosis for patients treated conservatively, limited number of study participants and missing values. Additionally, unique regional and internal hospital practices reduce the comparability of the collected datasets and transferability of the models, which, as we demonstrated, cannot be easily compensated for with model updating. Nonetheless, the observed distributions of parameters from both cohorts are clinically acceptable and display variability that is within expectations. Notably, our study allows to contrast the situatedness of a pediatric hospital against a general hospital where adult surgery and interdisciplinary surgical primary care are dominant. Lastly, the documented clinical, laboratory and ultrasound features are standardized, practical and cost-effective, enabling future analysis and comparison of our models on data from other institutions.

## 5 Conclusion

In this study, we performed an external validation of machine learning models for predicting the diagnosis, management and severity of pediatric appendicitis. When tested externally, the models exhibited lower predictive performance than on the original data. This was in part due to the shift in the prevalence of appendicitis cases we observed between the original and external datasets. Other possible reasons included intrinsic differences in patient demographics, clinical pathways, variations in referral practices and documentation standards for the two hospitals as well as the downstream effects of the COVID-19 pandemic. Such factors demonstrate the challenges of transferring predictive models between hospitals, which should always be done with care to avoid harmful fallout. As a potential remedy, we investigated model retraining; while it showed promise in restoring predictive performance, further research is necessary to determine the limitations of this approach, which we will explore in our future work. Additionally, we plan to investigate the possible design of the prospective evaluation and deployment of our predictive models. Specifically, we will look into defining the number of necessary blood tests and introducing standardized reporting guidelines for clinical examination and ultrasound findings.

## Data Availability Statement

The analyzed dataset in an anonymized form is available alongside the code in the following GitHub repository: https://github.com/i6092467/pediatric-appendicitis-ml-ext. Further inquiries can be directed to the corresponding authors.

## Ethics Statement

The studies involving humans were approved by the Ethics Committee of the University of Regensburg (18-1063-101, 18-1063-3-101). The studies were conducted in accordance with the local legislation and institutional requirements. In accordance with national legislation and institutional requirements, written informed consent was not required from participants or their legal guardians/next of kin for analysis of retrospective data. Informed consent was obtained from participants or their legal representatives for any follow-up investigation.

## Author Contributions

*Conceptualization:* Ričards Marcinkevičs, Patricia Reis Wolfertstetter.

*Writing – Original Draft:* Ričards Marcinkevičs, Kacper Sokol, Patricia Reis Wolfertstetter.

*Writing – Review & Editing:* Ričards Marcinkevičs, Kacper Sokol, Akhil Paulraj, Melinda A. Hilbert, Vivien Rimili, Sven Wellmann, Christian Knorr, Bertram Reingruber, Julia E. Vogt, Patricia Reis Wolfertstetter.

*Formal Analysis:* Ričards Marcinkevičs, Kacper Sokol, Akhil Paulraj.

*Data Curation:* Melinda A. Hilbert, Vivien Rimili, Patricia Reis Wolfertstetter.

*Investigation:* Ričards Marcinkevičs, Kacper Sokol, Akhil Paulraj, Melinda A. Hilbert, Vivien Rimili, Patricia Reis Wolfertstetter.

*Methodology:* Ričards Marcinkevičs, Kacper Sokol, Christian Knorr, Bertram Reingruber, Patricia Reis Wolfertstetter.

*Software:* Ričards Marcinkevičs, Kacper Sokol, Akhil Paulraj.

*Validation:* Ričards Marcinkevičs, Kacper Sokol, Akhil Paulraj.

*Project Administration:* Julia E. Vogt, Patricia Reis Wolfertstetter.

*Resources:* Julia E. Vogt.

*Supervision:* Sven Wellmann, Christian Knorr, Bertram Reingruber, Julia E. Vogt, Patricia Reis Wolfertstetter.

*Visualization:* Ričards Marcinkevičs, Kacper Sokol.

## Funding

The authors declare that no financial support was received for the research and/or publication of this article. The open access funding was provided by ETH Zurich.

## Conflict of Interest

The authors declare that the research was conducted in the absence of any commercial or financial relationships that could be construed as a potential conflict of interest.

## Generative AI Statement

The authors declare that no Generative AI was used in the creation of this manuscript.

## Footnotes

1 This dataset is available at https://github.com/i6092467/pediatric-appendicitis-ml.

## Notes

### Competing Interest Statement

The authors have declared no competing interest.

### Summary of Updates

New document layout for improved readability. Minor text revisions.

## References

[1] Amish Acharya, Sheraz R. Markar, Melody Ni, and George B. Hanna. 2016. Biomarkers of acute appendicitis: Systematic review and cost–benefit trade-off analysis. Surgical Endoscopy 31, 3 (2016), 1022–1031. doi:10.1007/s00464-016-5109-1

[2] Omer F Akmese, Gul Dogan, Hakan Kor, Hasan Erbay, and Emre Demir. 2020. The use of machine learning approaches for the diagnosis of acute appendicitis. Emergency Medicine International 2020, 1 (2020), 7306435. doi:10.1155/2020/7306435

[3] Pedro Roig Aparicio, Ričards Marcinkevičs, Patricia Reis Wolfertstetter, Sven Wellmann, Christian Knorr, and Julia E Vogt. 2021. Learning medical risk scores for pediatric appendicitis. In 2021 20th IEEE International Conference on Machine Learning and Applications (ICMLA). IEEE, 1507–1512. doi:10.1109/icmla52953.2021.00243

[4] Emrah Aydin, İnan Utku Türkmen, Gözde Namli, Çiğdem Öztürk, Ayşe B Esen, Y Nur Eray, Egemen Eroğlu, and Fatih Akova. 2020. A novel and simple machine learning algorithm for preoperative diagnosis of acute appendicitis in children. Pediatric Surgery International 36, (2020), 735–742. doi:10.1007/s00383-020-04655-7

[5] Roshanak Benabbas, Mark Hanna, Jay Shah, and Richard Sinert. 2017. Diagnostic Accuracy of History, Physical Examination, Laboratory Tests, and Point-of-care Ultrasound for Pediatric Acute Appendicitis in the Emergency Department: A Systematic Review and Meta-analysis. Academic Emergency Medicine 24, 5 (2017), 523–551. doi:10.1111/acem.13181

[6] Yoav Benjamini and Yosef Hochberg. 1995. Controlling the False Discovery Rate: A Practical and Powerful Approach to Multiple Testing. Journal of the Royal Statistical Society Series B: Statistical Methodology 57, 1 (1995), 289–300. doi:10.1111/j.2517-6161.1995.tb02031.x

[7] George S. Bethell, Tom Gosling, Clare M. Rees, Jonathan Sutcliffe, Nigel J. Hall, and CASCADE Study Collaborators and RIFT Study Collaborators. 2022. Impact of the COVID-19 pandemic on management and outcomes of children with appendicitis: The Children with AppendicitiS during the CoronAvirus panDEmic (CASCADE) study. Journal of Pediatric Surgery 57, 10 (2022), 380–385. doi:10.1016/j.jpedsurg.2022.03.029

[8] Aneel Bhangu, Kjetil Søreide, Salomone Di Saverio, Jeanette Hansson Assarsson, and Frederick Thurston Drake. 2015. Acute appendicitis: Modern understanding of pathogenesis, diagnosis, and management. The Lancet 386, 10000 (2015), 1278–1287. doi:10.1016/s0140-6736(15)00275-5

[9] William Bonadio, Peter Peloquin, Jared Brazg, Ilyssa Scheinbach, James Saunders, Chukwujekwu Okpalaji, and Peter Homel. 2015. Appendicitis in preschool aged children: Regression analysis of factors associated with perforation outcome. Journal of Pediatric Surgery 50, 9 (2015), 1569–1573. doi:10.1016/j.jpedsurg.2015.02.050

[10] Leo Breiman. 2001. Random Forests. Machine Learning 45, 1 (2001), 5–32. doi:10.1023/a:1010933404324

[11] Emily Decker, Agnes Ndzi, Simon Kenny, and Rachel Harwood. 2024. Systematic Review and Meta-analysis to Compare the Short- and Long-term Outcomes of Non-operative Management With Early Operative Management of Simple Appendicitis in Children After the COVID-19 Pandemic. Journal of Pediatric Surgery 59, 6 (2024), 1050–1057. doi:10.1016/j.jpedsurg.2023.12.021

[12] Louise Deleger, Holly Brodzinski, Haijun Zhai, Qi Li, Todd Lingren, Eric S Kirkendall, Evaline Alessandrini, and Imre Solti. 2013. Developing and evaluating an automated appendicitis risk stratification algorithm for pediatric patients in the emergency department. Journal of the American Medical Informatics Association 20, e2 (2013), e212–e220. doi:10.1136/amiajnl-2013-001962

[13] Salomone Di Saverio, Mauro Podda, Belinda De Simone, Marco Ceresoli, Goran Augustin, Alice Gori, Marja Boermeester, Massimo Sartelli, Federico Coccolini, Antonio Tarasconi, Nicola de’ Angelis Dieter G. Weber, Matti Tolonen, Arianna Birindelli, Walter Biffl, Ernest E. Moore, Michael Kelly, Kjetil Soreide, Jeffry Kashuk, Richard Ten Broek, Carlos Augusto Gomes, Michael Sugrue, Richard Justin Davies, Dimitrios Damaskos, Ari Leppäniemi, Andrew Kirkpatrick, Andrew B. Peitzman, Gustavo P. Fraga, Ronald V. Maier, Raul Coimbra, Massimo Chiarugi, Gabriele Sganga, Adolfo Pisanu, Gian Luigi de’ Angelis Edward Tan, Harry Van Goor, Francesco Pata, Isidoro Di Carlo, Osvaldo Chiara, Andrey Litvin, Fabio C. Campanile, Boris Sakakushev, Gia Tomadze, Zaza Demetrashvili, Rifat Latifi, Fakri Abu-Zidan, Oreste Romeo, Helmut Segovia-Lohse, Gianluca Baiocchi, David Costa, Sandro Rizoli, Zsolt J. Balogh, Cino Bendinelli, Thomas Scalea, Rao Ivatury, George Velmahos, Roland Andersson, Yoram Kluger, Luca Ansaloni, and Fausto Catena. 2020. Diagnosis and treatment of acute appendicitis: 2020 update of the WSES Jerusalem guidelines. World Journal of Emergency Surgery 15, 1 (2020). doi:10.1186/s13017-020-00306-3

[14] Jens Dingemann and Benno Ure. 2012. Imaging and the Use of Scores for the Diagnosis of Appendicitis in Children. European Journal of Pediatric Surgery 22, 03 (2012), 195–200. doi:10.1055/s-0032-1320017

[15] Alexandre Escribá, Anna Maria Gamell, Yolanda Fernández, Jose María Quintillá, and Carlos Luaces Cubells. 2011. Prospective Validation of Two Systems of Classification for the Diagnosis of Acute Appendicitis. Pediatric Emergency Care 27, 3 (2011), 165–169. doi:10.1097/pec.0b013e31820d6460

[16] Jerome H. Friedman. 2001. Greedy function approximation: A gradient boosting machine. The Annals of Statistics 29, 5 (2001). doi:10.1214/aos/1013203451

[17] Patrick Godau, Piotr Kalinowski, Evangelia Christodoulou, Annika Reinke, Minu Tizabi, Luciana Ferrer, Paul F. Jäger, and Lena Maier-Hein. 2023. Deployment of Image Analysis Algorithms Under Prevalence Shifts. In Medical Image Computing and Computer Assisted Intervention – MICCAI 2023. Springer, 389–399. doi:10.1007/978-3-031-43898-1_38

[18] Chung-Ho Hsieh, Ruey-Hwa Lu, Nai-Hsin Lee, Wen-Ta Chiu, Min-Huei Hsu, and Yu-Chuan Jack Li. 2011. Novel solutions for an old disease: Diagnosis of acute appendicitis with random forest, support vector machines, and artificial neural networks. Surgery 149, 1 (2011), 87–93. doi:10.1016/j.surg.2010.03.023

[19] George C. Koberlein, Andrew T. Trout, Cynthia K. Rigsby, Ramesh S. Iyer, Adina L. Alazraki, Sudha A. Anupindi, Dianna M.E. Bardo, Brandon P. Brown, Sherwin S. Chan, Tushar Chandra, Jonathan R. Dillman, Scott R. Dorfman, Richard A. Falcone, Matthew D. Garber, Madeline M. Joseph, Jie C. Nguyen, Nabile M. Safdar, and Boaz Karmazyn. 2019. ACR Appropriateness Criteria ® Suspected Appendicitis-Child. Journal of the American College of Radiology 16, 5 (2019), S252–S263. doi:10.1016/j.jacr.2019.02.022

[20] Ričards Marcinkevičs, Patricia Reis Wolfertstetter, Ugne Klimiene, Kieran Chin-Cheong, Alyssia Paschke, Julia Zerres, Markus Denzinger, David Niederberger, Sven Wellmann, Ece Ozkan, Christian Knorr, and Julia E. Vogt. 2024. Interpretable and intervenable ultrasonography-based machine learning models for pediatric appendicitis. Medical Image Analysis 91, (2024), 103042. doi:10.1016/j.media.2023.103042

[21] Ricards Marcinkevics, Patricia Reis Wolfertstetter, Sven Wellmann, Christian Knorr, and Julia E Vogt. 2021. Using machine learning to predict the diagnosis, management and severity of pediatric appendicitis. Frontiers in Pediatrics 9, (2021), 662183. doi:10.3389/fped.2021.662183

[22] Dmitri Nepogodiev, Richard JW Wilkin, Catherine J Bradshaw, Clare Skerritt, Alasdair Ball, Waaka Moni-Nwinia, Ruth Blanco-Colino, Priyesh Chauhan, Thomas M Drake, Matteo Frasson, et al. 2020. Appendicitis risk prediction models in children presenting with right iliac fossa pain (RIFT study): A prospective, multicentre validation study. The Lancet Child & Adolescent Health 4, 4 (2020), 271–280. doi:10.1016/S2352-4642(20)30006-7

[23] Go Ohba, Seiichi Hirobe, and Koji Komori. 2016. The Usefulness of Combined B Mode and Doppler Ultrasonography to Guide Treatment of Appendicitis. European Journal of Pediatric Surgery 26, 06 (2016), 533–536. doi:10.1055/s-0035-1570756

[24] H. C. Park, M. J. Kim, and B. H. Lee. 2017. Randomized clinical trial of antibiotic therapy for uncomplicated appendicitis. British Journal of Surgery 104, 13 (2017), 1785–1790. doi:10.1002/bjs.10660

[25] R Core Team. 2021. R: A Language and Environment for Statistical Computing. R Foundation for Statistical Computing, Vienna, Austria. https://www.R-project.org/

[26] Pranav Rajpurkar, Allison Park, Jeremy Irvin, Chris Chute, Michael Bereket, Domenico Mastrodicasa, Curtis P Langlotz, Matthew P Lungren, Andrew Y Ng, and Bhavik N Patel. 2020. AppendiXNet: Deep learning for diagnosis of appendicitis from a small dataset of CT exams using video pretraining. Scientific Reports 10, 1 (2020), 3958. doi:10.1038/s41598-020-61055-6

[27] Patricia Reis Wolfertstetter, John Blanford Ebert, Judith Barop, Markus Denzinger, Michael Kertai, Hans J. Schlitt, and Christian Knorr. 2024. Suspected Simple Appendicitis in Children: Should We Use a Nonoperative, Antibiotic-Free Approach? An Observational Study. Children 11, 3 (2024), 340. doi:10.3390/children11030340

[28] Josephine Reismann, Alessandro Romualdi, Natalie Kiss, Maximiliane I Minderjahn, Jim Kallarackal, Martina Schad, and Marc Reismann. 2019. Diagnosis and classification of pediatric acute appendicitis by artificial intelligence methods: An investigator-independent approach. PloS One 14, 9 (2019), e0222030. doi:10.1371/journal.pone.0222030

[29] Rebecca M. Rentea, Shawn D. St. Peter, and Charles L. Snyder. 2016. Pediatric appendicitis: State of the art review. Pediatric Surgery International 33, 3 (2016), 269–283. doi:10.1007/s00383-016-3990-2

[30] Robin Rey, Renato Gualtieri, Giorgio La Scala, and Klara Posfay Barbe. 2024. Artificial Intelligence in the Diagnosis and Management of Appendicitis in Pediatric Departments: A Systematic Review. European Journal of Pediatric Surgery 34, 05 (2024), 385–391. doi:10.1055/a-2257-5122

[31] Frank-Mattias Schäfer, Johannes Meyer, Stephan Kellnar, Jakob Warmbrunn, Tobias Schuster, Stefanie Simon, Thomas Meyer, Julia Platzer, Jochen Hubertus, Sigurd T. Seitz, Christian Knorr, and Maximilian Stehr. 2021. Increased Incidence of Perforated Appendicitis in Children During COVID-19 Pandemic in a Bavarian Multi-Center Study. Frontiers in Pediatrics 9 (2021). doi:10.3389/fped.2021.683607

[32] Kacper Sokol, James Fackler, and Julia E Vogt. 2025. Artificial intelligence should genuinely support clinical reasoning and decision making to bridge the translational gap. npj Digital Medicine 8, 1 (2025), 1–11. doi:10.1038/s41746-025-01725-9

[33] Carolin Stiel, Julia Elrod, Michaela Klinke, Jochen Herrmann, Carl-Martin Junge, Tarik Ghadban, Konrad Reinshagen, and Michael Boettcher. 2020. The modified Heidelberg and the AI appendicitis score are superior to current scores in predicting appendicitis in children: A two-center cohort study. Frontiers in Pediatrics 8, (2020), 592892. doi:10.3389/fped.2020.592892

[34] J. Svensson, N. Hall, S. Eaton, A. Pierro, and T. Wester. 2012. A Review of Conservative Treatment of Acute Appendicitis. European Journal of Pediatric Surgery 22, 03 (2012), 185–194. doi:10.1055/s-0032-1320014

[35] Jan F. Svensson, Barbora Patkova, Markus Almström, Hussein Naji, Nigel J. Hall, Simon Eaton, Agostino Pierro, and Tomas Wester. 2015. Nonoperative Treatment With Antibiotics Versus Surgery for Acute Nonperforated Appendicitis in Children: A Pilot Randomized Controlled Trial. Annals of Surgery 261, 1 (2015), 67–71. doi:10.1097/sla.0000000000000835

[36] Florien S. van Royen, Karel G.M. Moons, Geert-Jan Geersing, and Maarten van Smeden. 2022. Developing, validating, updating and judging the impact of prognostic models for respiratory diseases. European Respiratory Journal 60, 3 (2022), 2200250. doi:10.1183/13993003.00250-2022

[37] Jenna Wiens, Suchi Saria, Mark Sendak, Marzyeh Ghassemi, Vincent X Liu, Finale Doshi-Velez, Kenneth Jung, Katherine Heller, David Kale, Mohammed Saeed, Pilar N Ossorio, Sonoo Thadaney-Israni, and Anna Goldenberg. 2019. Do no harm: A roadmap for responsible machine learning for health care. Nature Medicine 25, 9 (2019), 1337–1340. doi:10.1038/s41591-019-0548-6

[38] Jianfu Xia, Zhifei Wang, Daqing Yang, Rizeng Li, Guoxi Liang, Huiling Chen, Ali Asghar Heidari, Hamza Turabieh, Majdi Mafarja, and Zhifang Pan. 2022. Performance optimization of support vector machine with oppositional grasshopper optimization for acute appendicitis diagnosis. Computers in Biology and Medicine 143, (2022), 105206. doi:10.1016/j.compbiomed.2021.105206

[39] Augusto Zani, Warwick J. Teague, Simon A. Clarke, Munther J. Haddad, Sanjeev Khurana, Thomas Tsang, and Ramesh M. Nataraja. 2017. Can common serum biomarkers predict complicated appendicitis in children? Pediatric Surgery International 33, 7 (2017), 799–805. doi:10.1007/s00383-017-4088-1

